# Plethysmometry: an automated cuff-based method for estimating ranges of systolic, diastolic and mean arterial pressures

**DOI:** 10.64898/2026.10.01.26364471

**Authors:** Naveen Gangadharan, Bowya Baskaran, A Manavalan, M Praveen Kumar, M Anandhakumar, Benjamin Jebaraj, Suresh Devasahayam, Sathya Subramani

## Abstract

**Background:** Conventional cuff based methods report single systolic and diastolic pressure values and depend either on Korotkoff sounds or on empirical oscillometric algorithms.

**Methods:** We developed a patented cuff based method, termed plethysmometry, that simultaneously records cuff pressure and finger photoplethysmograms from the cuffed and contralateral arms. The contralateral photoplethysmogram serves as an internal physiological reference for pulse amplitude. Following an initial pressure ramp, 16 cuff pressure plateaus are applied. Absolute and normalized pulse amplitudes in the cuffed arm are related to cuff pressure to estimate lower and upper systolic and diastolic pressures. Mean arterial pressure is estimated independently from cuff pressure oscillation amplitude. The estimations are done by automated algorithms developed in MATLAB.

**Results:** The method reports lower and upper systolic, diastolic and mean arterial pressure ranges (during the recording period), together with their average values. Average pulse pressure is also reported. Graphs are provided for reference to check the accuracy of reported pressures.

**Conclusion:** Plethysmometry provides an objective digital record of the cuff pressure and pulse traces and estimates systolic and diastolic pressure ranges from the pulse response of the individual subject to graded cuff pressure. It also reports mean arterial pressure. Comparison of plethysmometric systolic and diastolic pressures with sphygmomanometrically derived pressures and diagnostic agreement are reported in a companion paper.

## Introduction

Currently, two cuff-based methods are used clinically for blood pressure (BP) measurement: sphygmomanometry and oscillometry. Both employ an inflatable cuff encircling the arm, which can be pressurized, transmitting the pressure to the underlying artery. Under normal conditions, blood flow through arteries is laminar. When a stethoscope is placed over the brachial artery at the elbow, no sounds are heard. In sphygmomanometry, external pressure is exerted on the artery through a cuff around the arm. When the cuff pressure exceeds the true systolic pressure in the underlying artery, the artery collapses, arterial flow ceases and no sounds are heard. When cuff pressure is below the true diastolic pressure, laminar flow is undisturbed and no sounds are heard as stated earlier. When cuff pressure lies between systolic and diastolic levels, flow is no longer continuous but occurs in spurts, and tapping sounds called Korotkoff sounds, can then be auscultated over the antecubital fossa. As the cuff is deflated from a high pressure, the cuff pressure at the first appearance of these sounds indicates systolic pressure, and the cuff pressure at their disappearance indicates diastolic pressure.

The origin of the Korotkoff sounds is debated. The possibilities include intermittent, turbulent flow, and/or excursions of the arterial wall when it opens and closes. Yet another theory is that the sounds are not from the brachial artery, but due to shear waves in surrounding tissues during pulse propagation.

Baranger et al. used ultrafast ultrasound and concluded that Korotkoff sounds are shear vibrations transmitted through surrounding tissues rather than ordinary sound waves generated within the arterial lumen. [3] In that interpretation, the waves translate to audible sound when the stethoscope diaphragm is compressed by the tissue shear waves. However, the shear waves in the surrounding tissue must still be initiated by arterial wall motions.

Arterial wall motion depends on compliance, which is determined by both the intrinsic properties of the arterial wall and the degree of arterial distension. As arterial filling and consequently intraluminal pressure increase, transmural pressure and wall tension also increase, reducing arterial compliance. At cuff pressures between systolic and diastolic pressure, external pressure partly counterbalances intraluminal pressure, thereby reducing transmural pressure and increasing compliance.

In simpler terms, cuff compression places the artery at a lower transmural pressure, where it may be more compliant and undergo greater wall displacement with each pulse. This greater wall motion may generate stronger shear waves in the surrounding tissue and thereby contribute to Korotkoff sounds. If Korotkoff sounds depend on arterial wall motion and tissue shear-wave transmission, intrinsic arterial stiffness could influence both sound amplitude and the cuff pressures at which the sounds become audible or disappear.

Therefore, in stiffer arteries, reduced wall motion could make Korotkoff sounds fainter, potentially altering SBP and DBP estimates. This provides a plausible, although not fully established, mechanism by which arterial properties may contribute to auscultatory measurement error. Accurate cuff-based BP measurement also depends on procedural factors such as cuff size, arm support, positioning of the arm at heart level, adequate patient rest, and correct measurement technique. In auscultatory measurement, observer hearing and interpretation are additional sources of error, whereas for oscillometric measurement, use of a properly validated device is essential. These factors have been emphasized in recent scientific statements and systematic reviews. [1,2]

The consequences of auscultatory errors leading to misdiagnosis of hypertension cannot be overstated. Furthermore, because sphygmomanometry serves as the reference standard for validating new technologies, any inherent errors can propagate across the newer devices.

Validated automated devices have become the recommended method for hypertension diagnosis and management worldwide, because they eliminate many user errors. [1,18] The oscillometric method relies solely on cuff-pressure oscillations with each pulse to report pressures. In this method, cuff pressure is ramped up or down and cuff pressure recorded. The cuff pressure shows tiny oscillations whenever there is a pulse (see Figure 4). The cuff-pressure waveform is high-pass filtered to obtain the oscillations alone.

The cardinal principle of oscillometry is that oscillations have maximal amplitude when cuff pressure approximates mean arterial pressure (MAP), although exceptions occur. The most reliably determined parameter in oscillometry is the MAP, as identifying the cuff pressure at which maximum oscillation amplitude occurs is relatively straightforward.

The maximum oscillation amplitude (MOA) is determined and systolic and diastolic pressures are determined as cuff pressures above and below that of the MOA where the oscillation amplitude is a fixed ratio of the MOA. [4]

In the commonly described fixed-ratio method, systolic and diastolic pressures are assigned to cuff pressures where the oscillation amplitude reaches empirically derived fractions of the maximum oscillation amplitude. Sharman *et al*. cite approximate coefficients of 0.50 for systolic and 0.70 for diastolic pressure, designed to correspond with auscultatory sphygmomanometric values, while emphasizing that optimal coefficients vary considerably among devices and individuals. [4] Chandrasekhar *et al*. explicitly describe these coefficients as population-based fractions of maximum oscillation amplitude. Their analysis demonstrates that the true ratios vary substantially with arterial compliance and pulse pressure, so errors in the assumed ratio may translate into appreciable BP error. [5]

Alvarado Alvarez *et al*. [6] systematically evaluated fixed-amplitude ratios by comparing oscillometric BP estimates with corresponding auscultatory reference pressures in 219 measurements obtained from 73 healthy volunteers. They sought to identify combinations of amplitude ratios and oscillometric-envelope representations that would satisfy the numerical accuracy limits commonly used in standard BP-device validation—namely, a mean device–reference error of ≤5 mmHg and a standard deviation of error <8 mmHg relative to the auscultatory reference.

Depending on the representation of the oscillometric envelope, systolic amplitude ratios ranging from 0.44 to 0.74 and diastolic ratios ranging from 0.51 to 0.85 satisfied these population-level accuracy limits. However, when optimal ratios and envelope representations were selected on the basis of the population mean, the highest proportions of participants with BP estimates within the specified limits were only 72.6% for systolic pressure and 69.9% for diastolic pressure. The authors therefore concluded that a considerable proportion of the population continued to have unreliable estimates.

These findings illustrate that multiple combinations of fixed ratios and waveform-envelope processing can produce acceptable group-level mean error and standard deviation, while appreciable errors may persist in individual participants. Published fixed-ratio methods therefore depend on empirically derived group-level relationships rather than exclusively on the pressure–oscillation-amplitude response of the individual being examined. Commercial devices may employ more complex proprietary algorithms; however, their derivation datasets, coefficients and processing rules are generally not publicly disclosed.

Several investigators have compared cuff-based BP estimates with invasive brachial and aortic pressures. Invasive measurements demonstrate important pressure differences along the arterial tree. Brachial systolic pressure is generally higher than aortic systolic pressure because of peripheral pressure amplification, whereas diastolic pressure differs less between central and peripheral sites. Cuff BP was historically intended to approximate central aortic pressure, because the heart and major target organs are exposed more directly to central than to peripheral arterial pressure. [7,8]

In their systematic review and meta-analysis, Picone *et al*. found substantial disagreement between cuff-based and invasive measurements, with marked variation among individuals. Cuff methods tended to underestimate invasive brachial systolic pressure and overestimate brachial diastolic pressure. When compared with intra-aortic pressure, cuff systolic pressure showed both over- and underestimation across studies, whereas diastolic pressure was more consistently overestimated. [7] These findings indicate that cuff pressure cannot be assumed to correspond uniformly either to invasive brachial pressure or to central aortic pressure.

The magnitude and direction of this error may also vary with physiological characteristics. Bui *et al*. reported that age influences the relationship between cuff and aortic pressure: oscillometric devices may overestimate aortic systolic pressure in younger adults and underestimate it in older adults, while diastolic pressure tends to be overestimated, with the discrepancy increasing with age. [8] More recently, Picone *et al*. showed that oscillometric MAP itself may differ clinically from invasive aortic MAP and that MAP error may contribute to subsequent SBP and DBP estimation error. [9]

These discrepancies are clinically important because numerical disagreement becomes particularly consequential when it changes an individual’s BP classification. Diagnostic thresholds also differ among guidelines: many international recommendations continue to define office hypertension as SBP ≥140 mmHg and/or DBP ≥90 mmHg [10,11], whereas the ACC/AHA guideline used in the United States defines hypertension from an average SBP ≥130 mmHg or DBP ≥80 mmHg. Picone *et al*. reported that cuff readings below 120/80 mmHg or above 160/100 mmHg provided greater confidence in classification, whereas classification was less reliable in the intermediate range of approximately 120/80–160/100 mmHg. [7] Thus, acceptable mean agreement at the group level does not necessarily ensure accurate classification in individuals, particularly when measured pressures lie near a diagnostic threshold.

A further under-recognized feature of BP is its short-term variability over minutes, driven by Traube (respiratory) and Mayer (low-frequency) waves. From intra-arterial recordings in 51 hemodynamically stable ICU patients, we previously reported variability of 21 ± 9 mmHg for systolic and 14 ± 5 mmHg for diastolic pressure over a 10-minute period [12]. Single point estimates of this widely varying parameter can lead to misdiagnosis as well. Blood pressure variability has also been associated with target organ damage independent of mean BP in several studies and reviews. [13-15] Few clinically practical non-invasive devices provide an estimate of short-term BP variability during a single supervised recording. The above considerations underscore the need for alternative approaches to measuring BP.

We have developed a cuff-based method for estimating ranges of systolic and diastolic BP from finger pulse-volume changes measured using photo-plethysmography (PPG). The systolic pressure range is estimated from the cuff-pressure interval over which the distal pulse transitions from complete pulse presence to complete pulse absence. The diastolic pressure range is estimated from the cuff-pressure region over which pulse-volume approaches the unpressurized state. The method is hereafter referred to as ‘Plethysmometry’. PPG is increasingly being explored as a source of BP-related vascular information, but most contemporary PPG-based BP methods are cuff-less or model-based and remain limited by the need for calibration against a cuff-based reference method, as well as by challenges in generalizability and validation. [16,17]

Plethysmometry differs from these approaches by using controlled cuff-pressure plateaus to directly relate pulse-volume changes to cuff-pressure. This allows the pressure estimates to be derived from the individual’s cuff-pressure–pulse-volume response rather than from a fixed population-derived amplitude ratio.

PPG amplitude at a given cuff pressure is not uniform but varies due to variations in diastolic and/or systolic pressures. Maintaining cuff pressure at a plateau for 10 seconds allows capturing variations in PPG amplitude at a given cuff pressure. The relationship of varying PPG amplitudes to cuff pressure allows determination of the range of systolic and diastolic pressures.

Unlike conventional cuff methods that report a single systolic and diastolic pressure, plethysmometry estimates the range of systolic, diastolic, and mean arterial pressures during the measurement interval. The average of the range provides a summary value that can be compared with conventional single-value reports, while retaining information on the observed range. The method reports MAP independently by assessing cuff-pressure oscillation amplitudes at different cuff pressures.

This first companion paper describes the physiological basis, instrumentation, recording protocol and analytical approach of plethysmometry. The accompanying second paper evaluates agreement with carefully obtained reference auscultatory sphygmomanometry and explores diagnostic classification for hypertension.

## Methods Plethysmometric system

The patented plethysmometric system consists of an inflatable upper-arm cuff, a cuff-pressure transducer, a digitally controlled pump, and two photoplethysmographic sensors. One photoplethysmographic sensor is placed on a finger of the cuffed arm, while a second sensor is placed on the corresponding finger of the contralateral arm and serves as a reference signal.

Cuff pressure and photoplethysmographic waveforms from the cuffed and uncuffed arms are acquired simultaneously through three recording channels at a sampling frequency of 500 samples per second.

The recorded signals are subsequently processed to estimate the lower and upper limits of systolic, diastolic, and mean arterial pressures during the recording period.

### Participant preparation

Participants were seated comfortably in an armless chair with back support. Mid-arm circumference was measured, and an appropriately-sized cuff was placed on the upper arm. Photoplethysmographic sensors were attached to corresponding fingers of both hands. The forearms and hands were supported on the thighs, with the palms facing upward, and participants were instructed to remain still during recording.

### Cuff-pressure protocol

The plethysmometric recording comprised an initial ramp phase followed by a stepped-pressure phase. During the ramp phase, cuff pressure was increased continuously until the pulse in the cuffed arm disappeared. The cuff pressure at which pulse disappearance occurred was used as an approximate systolic-pressure estimate.

For the stepped-pressure phase, the pressure range from 30 mmHg to approximately 30 mmHg above the preliminary systolic estimate was divided into 16 levels. In the current protocol, these levels are applied in ascending order, beginning at 30 mmHg. Each pressure level is maintained for approximately 10 seconds and followed by complete cuff deflation for a further 10 seconds. The companion comparison study was conducted using an earlier version of the protocol in which the same pressure levels were applied in descending order. The analytical method applied to each pressure plateau did not change.

Cuff pressure and photoplethysmographic signals from both arms were recorded continuously throughout the procedure. The complete ramp-and-step recording lasted approximately six minutes.

The 10-second duration of each plateau was selected to permit recording of several cardiac cycles and, in most participants, to include changes occurring over at least one respiratory cycle.

At cuff pressures above systolic pressure, pulses in the cuffed arm were absent. At pressures below diastolic pressure, cuffed-arm pulse amplitude approached that recorded in the uncompressed state. Between these pressure levels, the cuffed-arm pulse remained detectable but was attenuated.

### Signal processing and pulse-amplitude measurement

The photoplethysmographic signals were filtered using a passband of 0.5 to 10 Hz. At each cuff-pressure plateau, pulse peaks and the preceding troughs were identified in both the cuffed-arm and reference-arm photoplethysmograms. Pulse amplitude was calculated as the difference between each pulse peak and its preceding trough.

For every pulse, the cuffed-arm pulse amplitude was analysed both as an absolute value and after normalization to the simultaneously recorded pulse amplitude in the contralateral reference arm.

Normalization was used to reduce the effect of systemic fluctuations in pulse amplitude unrelated to cuff compression, including respiratory and vasomotor changes.

For each pressure plateau, pulse amplitudes were related to the corresponding cuff pressure. The relationship between cuff pressure and cuffed-arm pulse amplitude was typically sigmoid.

The pulsatile component of cuff pressure at each plateau was separately extracted to quantify cuff-pressure oscillation amplitude.

### Estimation of systolic pressure

At cuff pressures below systolic pressure, some or all pulses remained detectable in the cuffed arm. At cuff pressures above systolic pressure, pulses were absent. The lower systolic limit was identified from the transition between a pressure plateau at which all pulses were present and the next pressure level at which one or more pulses were absent. The upper systolic limit was identified from the transition to the pressure level at which all cuffed-arm pulses were absent. Accordingly, systolic pressure was reported as a range rather than a single value.

### Estimation of diastolic pressure

At cuff pressures below diastolic pressure, pulse amplitude approached the uncompressed or reference-adjusted amplitude. At pressures above diastolic pressure, pulse amplitude progressively declined.

The algorithm considered both absolute and normalized pulse amplitudes, variability within each pressure plateau, and their relation to mean arterial pressure to identify the lower and upper limits of diastolic pressure.

### Estimation of mean arterial pressure

Mean arterial pressure was estimated independently from cuff-pressure oscillations. The pulsatile component of cuff pressure was extracted at each plateau, and the amplitude of the cuff-pressure oscillations was related to mean plateau pressure. The cuff pressure associated with the maximum oscillation amplitude was taken as the principal estimate of mean arterial pressure. A lower and upper MAP limit was reported when the maximal oscillatory amplitudes extended across more than one adjacent pressure level.

### Algorithm development and application

The analytical algorithm was developed from recurring PPG patterns observed in more than 1,000 recordings from an ongoing study. It incorporates the presence or absence of cuffed-arm pulses, absolute and normalized pulse amplitudes, within-plateau variability, sigmoid transition regions, and cuff-pressure oscillation amplitude. It was deterministic and rule-based, with no machine-learning component. Reference BP values were not used to derive, calibrate, or optimize the pressure-estimation rules. The algorithmic rules and parameter settings were finalized before the reference sphygmomanometric values from the 96-participant comparison cohort reported in the companion paper were examined. The cohort therefore provided an evaluation of the finalized pressure-estimation algorithm by comparison with reference values that had not been used during algorithm development.

For each recording, the algorithm reported:

- lower and upper systolic pressure;
- lower and upper diastolic pressure;
- lower and upper mean arterial pressure.

Average systolic, diastolic, and mean arterial pressures were calculated as the average of the lower and upper estimates. The range (in the duration of the recording) for each pressure parameter was defined as the difference between its upper and lower estimates. The physiological and clinical significance of this measure as an index of ultrashort-term BP variability requires further evaluation.

### Determination of systolic and diastolic pressures

Expanding the time axis shows absence of pulses in the plateau between 270 and 300 seconds, when cuff pressure was at 121 mmHg, and at all cuff pressures above 121 mmHg (Figure 2a). The highest systolic pressure in this subject is therefore 121 mmHg. All pulses are present, albeit at reduced amplitude, in the cuff pressure plateau between 240 and 260 seconds (average pressure = 112 mmHg).

**Figure 1:**
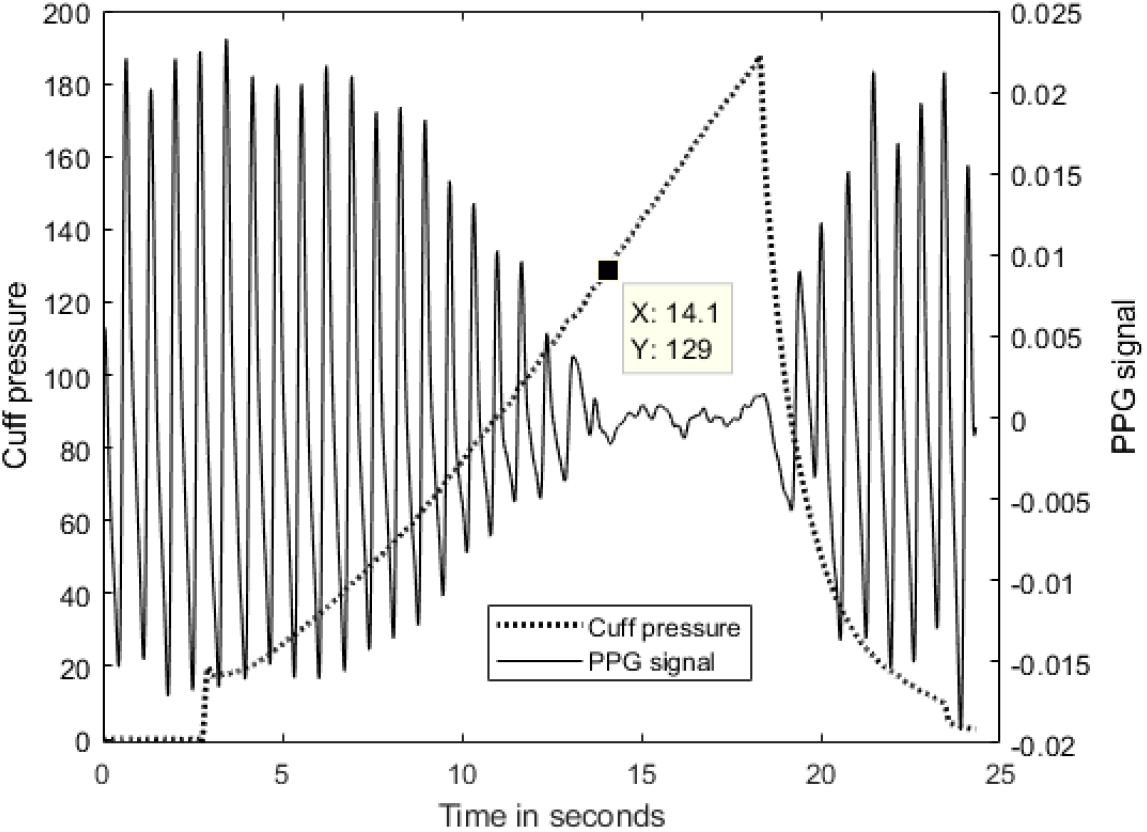
Ramp protocol for rough estimate of systolic pressure.

**Figure 2:**
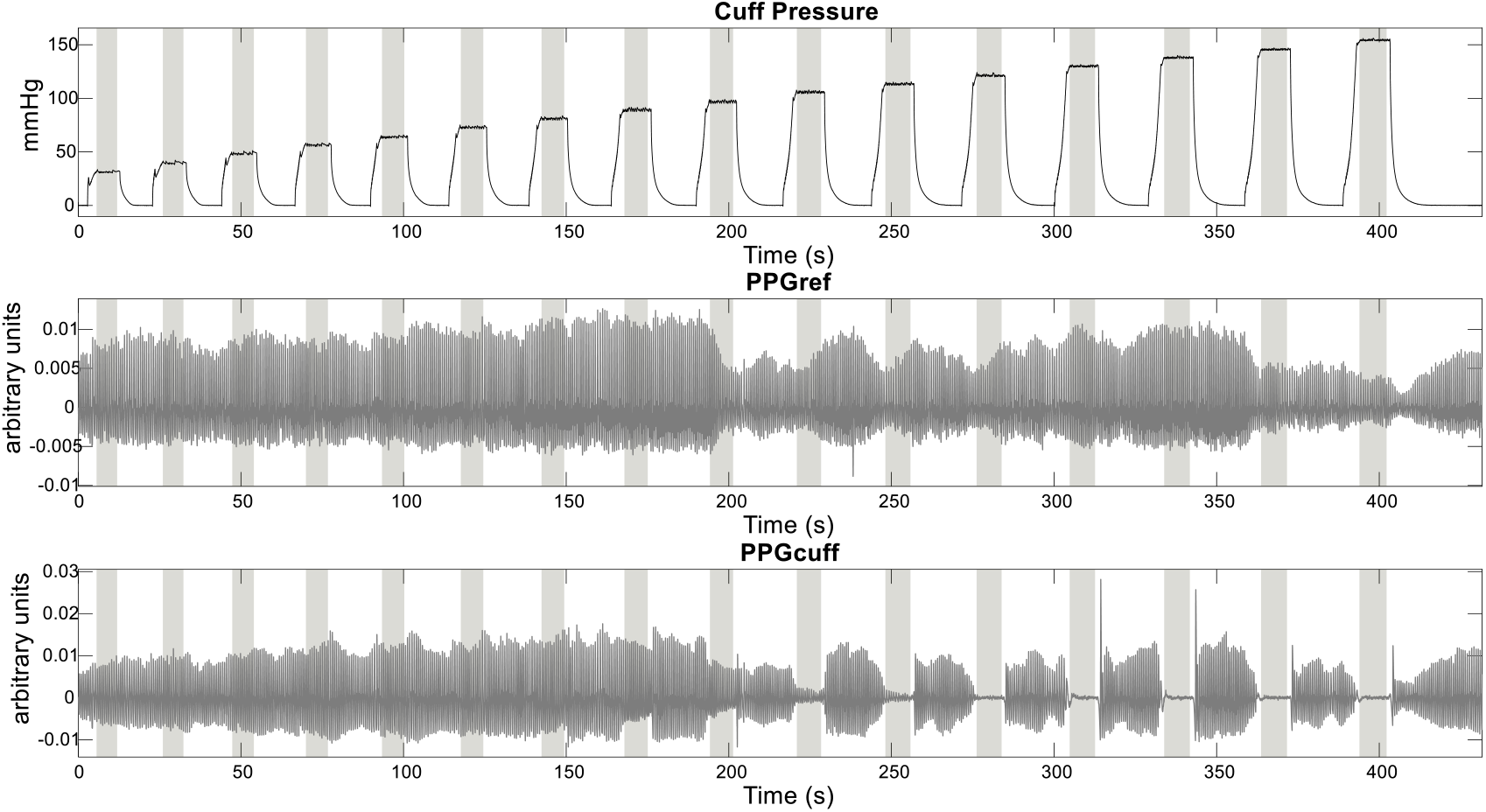
Cuff pressure, PPG from the uncuffed arm and PPG from the cuffed arm acquired simultaneously:

**Figure 2a:**
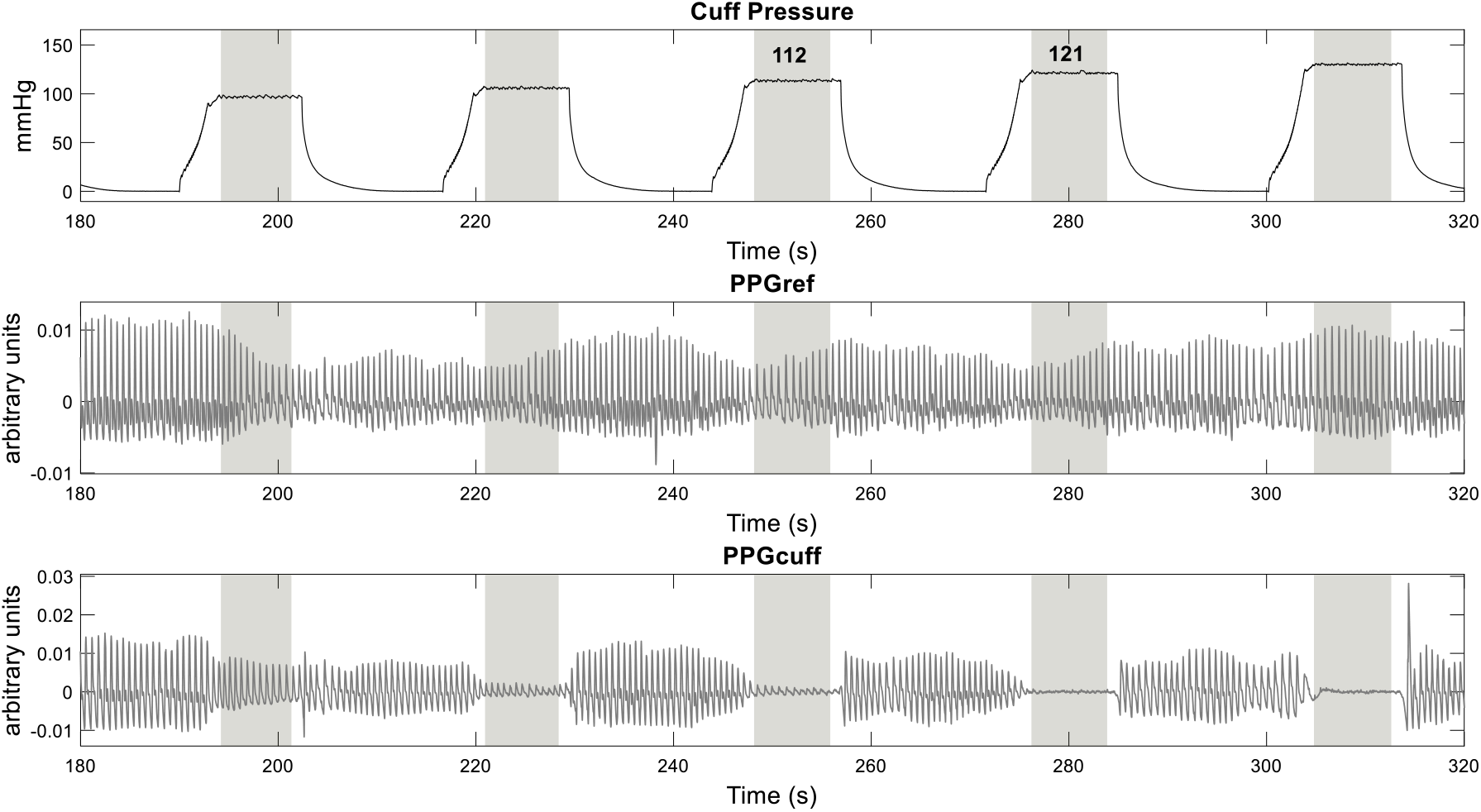
At a cuff pressure of 121 mmHg, pulses are absent in the cuffed arm.

The cuff pressure at which at least one pulse is absent is above the lower systolic pressure, and the cuff pressure at which all pulses are absent is above the highest systolic pressure in that individual. The lower and upper systolic estimates were derived from the adjacent cuff-pressure levels spanning the transition from complete pulse presence to complete pulse absence. Thus, the reported systolic range represents the pressure interval within which the pulse-occlusion transition occurred during the recording. In the example above, all pulses are present in the plateau between 240 and 260 seconds (113 mmHg) while all are absent in the next plateau (121 mmHg). The algorithm reported the systolic pressure range as 114 -121 mmHg.

Diastolic pressure is taken as the maximum cuff pressure beyond which pulse amplitude is less than normal. It is difficult to determine where this happens from the raw tracing. Plotting the peaks and troughs of the pulses during the plateau and determining the amplitude of each pulse allows plotting of Figure 3.

**Figure 3:**
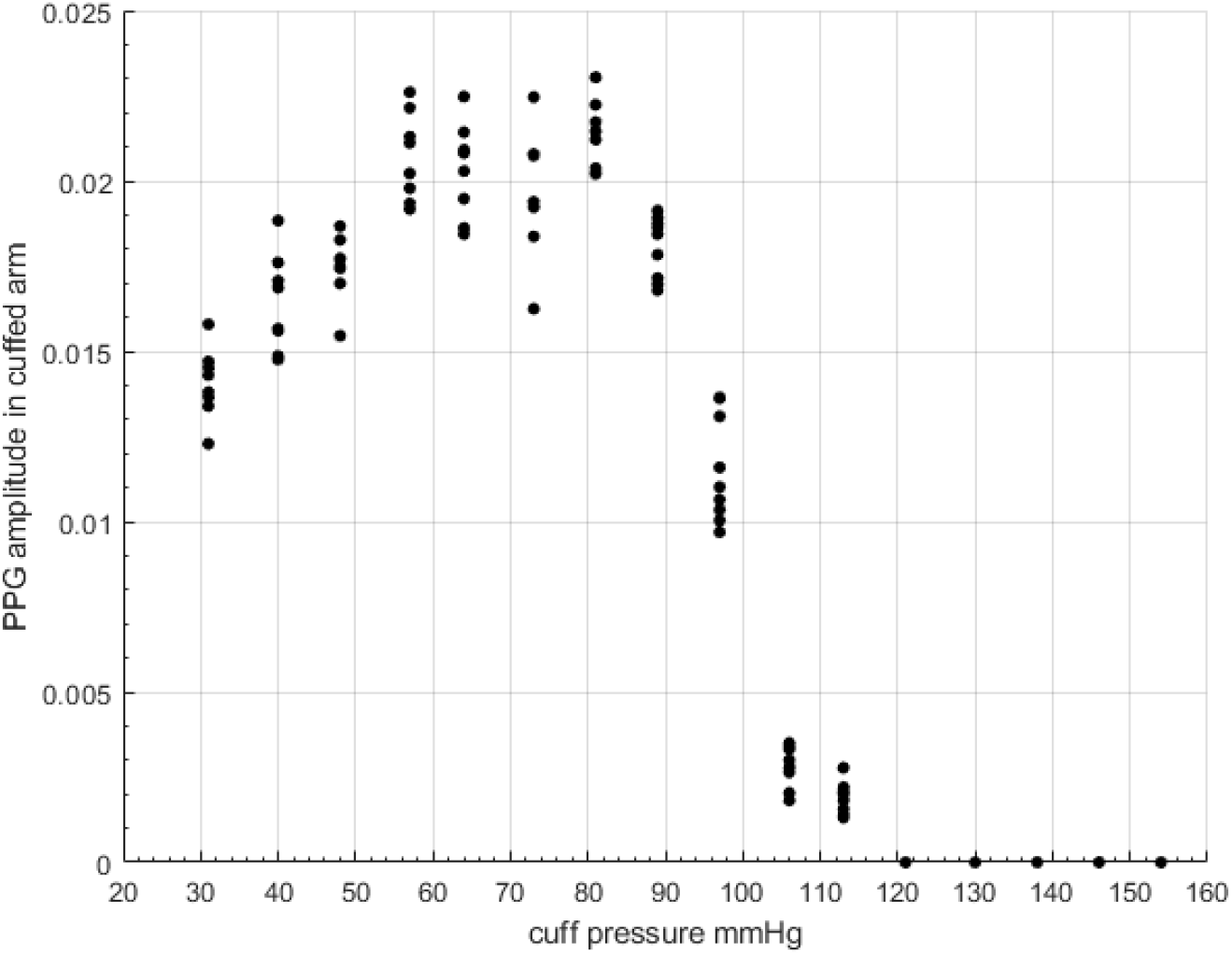
Relationship between cuff pressure and cuffed-arm PPG pulse amplitude. Each point represents the peak-to-preceding-trough amplitude of an individual pulse recorded during a cuff-pressure plateau. Pulse amplitude remains relatively preserved at lower cuff pressures and declines through the transition region as cuff pressure increases.

It can be seen that PPG amplitude begins to decline after 80 mmHg. As PPG amplitude can vary biologically in addition to changes determined by cuff pressure, cuff PPG amplitude is normalized to that of the reference arm, and both actual and normalized amplitudes are considered in the analytical algorithm that the device employs. Normalization is intended to reduce the contribution of systemic fluctuations in pulse amplitude unrelated to cuff compression.

As shown in figure 3, pulse amplitude declines at cuff pressures higher than 80 mmHg. The crude diastolic pressure estimate is around 80 mmHg. The algorithm considers pulse-amplitude variability, absolute and reference-normalized amplitudes, and their relationship to the independently estimated MAP when determining the lower and upper diastolic estimates.

### Determination of mean arterial pressure

MAP is determined independently as the cuff pressure at which the amplitude of the cuff-pressure oscillations is maximal. Figure 4 demonstrates oscillations in cuff pressure during a plateau.

**Figure 4:**
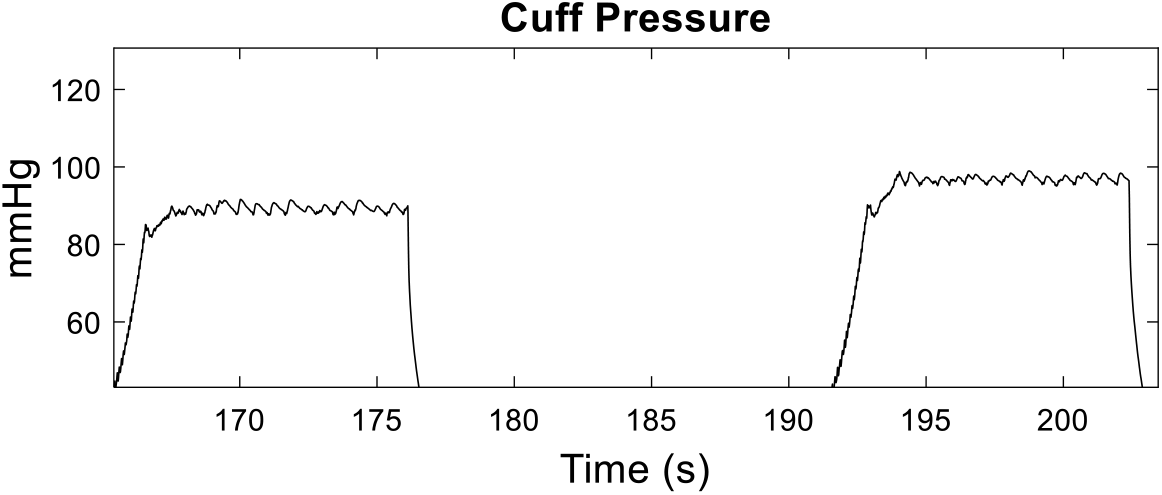
Cuff-pressure oscillations during a pulse.

The core theory on which the oscillometric method of BP measurement is based is that cuff-pressure oscillations have maximal amplitude when cuff pressure is at the mean arterial pressure. The maximal oscillations in this subject occur at about 90 mmHg cuff pressure (Figure 5).

**Figure 5:**
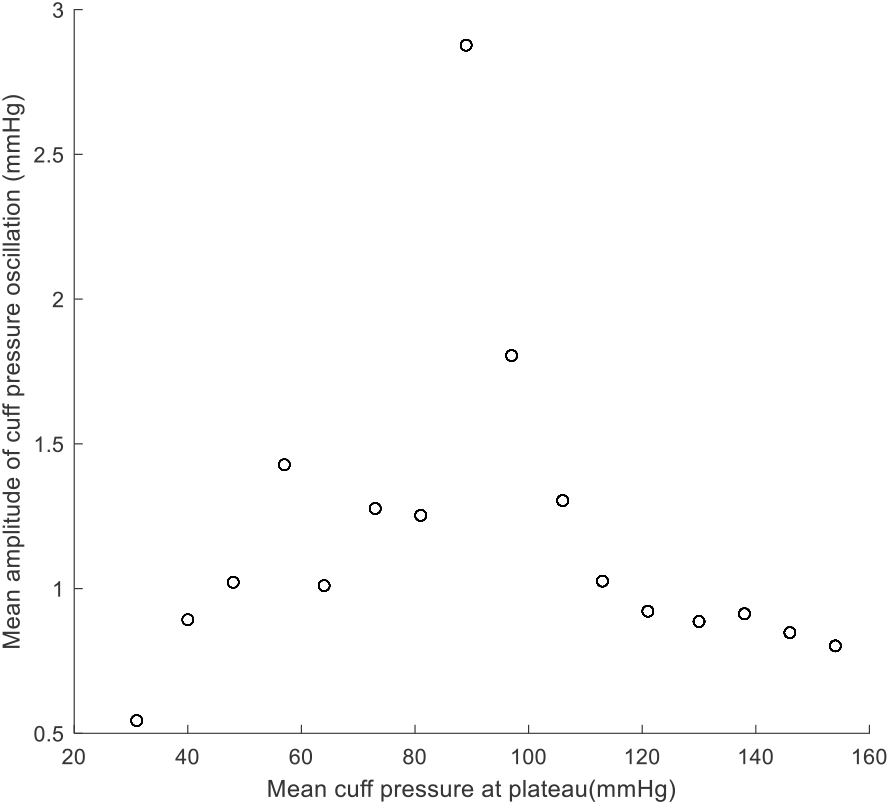
Average amplitude of cuff-pressure oscillations during a plateau plotted against mean plateau pressure.

The report of the algorithm for the above subject is as follows (all pressure values in mmHg).

| Lower systolic | Upper systolic | Lower diastolic | Upper diastolic | Lower MAP | Upper MAP |
| --- | --- | --- | --- | --- | --- |
| 114 | 121 | 75 | 79 | 89 | 90 |

The average (mid-point) pressures which may be compared with standard reports would be 118/77 mmHg (systolic/diastolic) with the mean arterial pressure being 90 mmHg. The systolic, diastolic and MAP ranges (in the duration of the recording) in this example are 7, 4 and 1 mmHg, respectively.

## Discussion

As plethysmometry relates cuff pressure to pulse-volume, the technology is inherently different from both sphygmomanometry and oscillometry for determination of systolic and diastolic pressures. It also uses oscillometric signals to determine mean arterial pressure. It provides the range of systolic, diastolic and mean arterial pressures. The average of the reported range provides a summary value that can be compared with conventional single-value reports, while the full range retains information on variation during the recording.

Conventional devices generally reduce the BP information acquired during a measurement cycle to a single reported systolic and diastolic value. Longer-term BP variability is therefore assessed through repeated measurements, as in ABPM or home monitoring.

Blood pressure variability itself is gaining importance as a risk predictor for target organ damage, independent of mean BP. In untreated hypertensives, beat-to-beat systolic pressure variability has been reported to relate to organ damage, and ambulatory systolic BP variability correlates with subclinical target-organ damage independent of mean BP. [12,13] Recent reviews and expert documents have also emphasized that BPV may provide additional cardiovascular risk information beyond average BP. [14] Few clinically practical non-invasive methods provide both pressure estimates and a measure of short-term variation within the recording session. The plethysmometric method provides a pressure range (in the duration of the recording) derived from PPG amplitude variation at controlled cuff-pressures. Whether this range provides a valid measure of ultrashort-term BP variability requires further investigation.

The advantages of plethysmometry over sphygmomanometry may be appreciated if the well-known limitations of sphygmomanometry are reiterated. The limitations of Korotkoff sounds themselves have been discussed in the introduction. In addition, sphygmomanometry is a subjective measurement; its accuracy depends on the hearing ability of the examiner and ambient noise levels. It requires trained personnel and careful attention to the fast-changing mercury level, or the pointer in aneroid devices.

Recent systematic reviews have highlighted multiple sources of inaccuracy in BP readings obtained in clinical settings. [2]

Plethysmometry can be done in noisy outpatient environments. Plethysmometry retains inspectable digital cuff-pressure and PPG traces from which the basis of the reported estimates can be reviewed; conventional commercial oscillometric devices generally do not expose the underlying waveform-based determination to the operator.

As discussed earlier, commonly described fixed-ratio oscillometric methods use empirically derived population-level relationships between oscillation amplitude and reference sphygmomanometric systolic and diastolic pressures. The optimal ratios vary among devices and individuals and are influenced by arterial properties and waveform-envelope processing. [4–6] Commercial algorithms may be more complex, but their coefficients and processing rules are generally proprietary.

The maximum oscillation amplitude is very clear in Figure 5, and the oscillation amplitudes at cuff pressures on either side of maximum are very low. Such a pattern gives a clear estimate of MAP at least. However, such clean records of MOA are not always obtained. In some individuals, oscillations may remain high beyond MAP, making the estimation of systolic pressure by fixed-ratio methods less transparent. Recent work showing device-specific MAP errors and the dependence of SBP/DBP error on MAP error supports the need for improved automated methods. [9]

A further conceptual difference is that plethysmometric estimation of systolic and diastolic pressures does not depend on population-derived fixed ratios. Plethysmometry derives systolic and diastolic pressure ranges from the relationship between cuff-pressure and the subject’s own pulse-volume. The estimate is therefore based on the individual pressure–pulse-volume response recorded during that examination, rather than on a generalized population ratio. Population data are still required for deriving diagnostic classification thresholds, but not for the primary physiological estimation of the pressure range itself. Also, unlike single-sensor approaches, the reference arm provides an internal physiological control that compensates for systemic changes occurring during measurement.

The challenge in incorporating plethysmometry into routine clinical practice will be the derivation of classification thresholds for hypertension diagnosis. There are three possible approaches to deriving such thresholds: (1) the ‘distribution-based approach,’ which uses a certain percentile of the BP distribution in the population as a threshold; (2) the ‘reference-comparison-based approach, including regression-or ROC-derived thresholds,’ which identifies plethysmometric systolic and diastolic BP levels that correspond to specific levels obtained from oscillometry or sphygmomanometry (e.g., 130/85, 140/90); and (3) the ‘outcome-based approach,’ which identifies a plethysmometric threshold that has the same risk for a CVD outcome as conventional thresholds (e.g., 140/90 mmHg). While it is best to have an outcome-derived approach for threshold identification, a distribution and/or regression-derived approach can be employed initially until outcomes are assessed in long-term cohorts. These ideas are considered in the adjoining paper.

## Conclusion

Plethysmometry is a cuff-based approach that relates graded external pressure to the individual’s pulse-volume response. It provides lower and upper estimates of systolic, diastolic and mean arterial pressure. Average values of systolic and diastolic pressure ranges can be used for comparison with conventional BP reports. The method also provides inspectable digital cuff-pressure and photoplethysmogram traces.

Systolic and diastolic estimation is based on the individual pressure–pulse-volume relationship rather than fixed oscillometric amplitude ratios, while MAP is derived independently from cuff-pressure oscillations. Further studies are required to establish the reproducibility and clinical significance of the reported pressure ranges and to validate diagnostic thresholds in independent populations.

## Data Availability

The datasets generated during and/or analyzed during the current study are available from the corresponding author on reasonable request.

## ACKNOWLEDGEMENTS

The authors thank the Dept. of Physiology, Christian Medical College, Vellore for enabling the study and data collection.

## FUNDING

The study was funded by Department of Biotechnology (DBT), Government of India and Indian Council of Medical Research.

## ETHICS

This study was approved by the Institutional Review Board of Christian Medical College, Vellore, IRB no: 14388, 22/12/2021

## CONFLICT OF INTEREST

The authors declare no competing interests.

## Notes

### Competing Interest Statement

The authors have declared no competing interest.

### Author Declarations

Institutional Review Board of Christian Medical College, Vellore, Tamilnadu, India

